# Hemoglobin specific volume width might promote the prevalence and poor long-term prognosis of American cardiovascular disease adult patients: the NHANES 1999-2020

**DOI:** 10.1101/2023.09.07.23295178

**Authors:** Lin Zhang, Jun Chang, Kaiyue Wang, Ying Zhou, Liming Wang, Lele Zhang, Xuemei Zhang, Zhifei Fu, Lifeng Han, Xiumei Gao

## Abstract

**Background:** Cardiovascular disease (CVD) patients are always accompanied by erythrocyte dysfunction. However, current erythrocyte-related indicators can’t explain the prevalence and long-term prognosis of CVD. Therefore, hemoglobin specific volume width (HSW) was first created to explain this phenomenon.

**Methods:** HSW’s quartiles were determined with Q1 [1.88,3.64] cL/g, Q2 (3.64,3.85] cL/g, Q3 (3.85,4.11] cL/g, and Q4 (4.11,11.74] cL/g. Among the 57,894 participants derived from National Health and Nutrition Examination Survey (NMAHES), 5,623 CVD patients had a whole following time (median 79 months). Logistic regression, Cox regression, Kaplan–Meier curves, and restricted cubic spline were utilized for analysis.

**Results:** Among the non-CVD (*n*=51,243) and CVD (*n*=6,651) individuals, the percentages of CVD in quartiles of HSW (Q1, Q2, Q3, and Q4) were 4.95%, 7.11%, 10.51%, and 16.14% (*P* < 0.0001). Adjusted odds ratio (OR) was still significant, 1.46 (95% CI 1.33, 1.60). Processing Q1 as reference, the adjusted ORs for Q3 and Q4 were 1.19(95% CI 1.05,1.35) and 1.54 (95% CI 1.36,1.73). Females had a higher risk (OR 1.49, 95% CI 1.32, 1.68) of CVD than males with increasing HSW. Diabetes mellitus (DM) patients had the highest risk of CVD (OR 1.54, 95% CI 1.30,1.84) than preDM or healthy individuals. Among dead (*n*=2,517) and alive CVD (*n*=3,106) patients, the adjusted HSW hazard ratio (HR) was 1.70 (95% CI 1.48,1.95). An inverse L-curve with an inflexion point of 3.96 was observed between HR and CVD. CVD patients with HSW > 3.96 had a poor prognosis than HSW <= 3.96.

**Conclusions:** HSW is a novel risk for CVD prevalence and long-term prognosis. Females and DM patients have a higher risk of CVD when HSW increases.

## 1. Introduction

Cardiovascular disease (CVD) is the primary cause of mortality ^1,2^, triggering poor outcomes, dramatic mortality, and high rehospitalization rates worldwide ^3^. Many red blood cells (RBC) related clinical indicators ^4–6^ were associated with CVD’s development or even mortality. For example, red blood distribution width (RDW) ^7,8^ and hemoglobin (Hb) were verified as independent predictors of CVD mortality. However, the hazard ratios (HR) of RDW and Hb were both too low, only 1.13 ^9^ or 0.8 ^10^, respectively. That is, although RDW and Hb independently predicted the prognosis of patients with CVD, the predictive values were not high. Are there other RBC-related indicators that can better predict the prognosis of patients with CVD?

Hemoglobin specific volume width (HSW), first proposed in this work, is a novel RBC-related measure that mirrors the volume fluctuation of RBC corresponding to every unit of hemoglobin. HSW is similar but different to RDW, a variation in RBC volume ^11^. But HSW emphasizes the variability volume fluctuation of RBC corresponding to every unit of hemoglobin. However, previous research work had not focused on HSW in any disease. And the calculation strategy of HSW was developed for the first time in this work. We hypothesized that there was a specific association between HSW and CVD, so relevant experiments were designed to verify it.

In summary, this work focuses on the relationship between HSW and CVD patients and further investigates the correlation between CVD prevalence and long-term prognosis in HSW. Three objectives are in this study: i) analyze the prevalence between HSW in non-CVD and CVD patients; ii) explore the optimal value of HSW to define the prognosis of CVD; iii) analyze the prognosis of CVD patients at various levels of HSW.

## 2. Methods

### 2.1 Calculation of HSW

As the inverse of density, the specific volume describes the volume per unit mass. It is widely applied in muscle testing ^12^ and food ^13–15^ but never in hemoglobin. For example, the muscle specific volume illustrates the skeletal muscle volume in different individuals ^12^. In the food field, the bread specific volume width indicated the fluctuation of the expanded bread volume with the same mass ^16,17^ at various conditions. In chemistry, liquid specific volumes emphasize the volume fluctuation ^18^ with the same mass at different temperatures.

Similarly, the HSW can reflect fluctuation in the volume of RBC with the same hemoglobin mass. Previous research had not focused on the HSW in any disease, and we first proposed it. So how to measure or calculate the HSW?

As the name implies, the mean corpuscular haemoglobin concentration (MCHC) ^19^ is the mean value of hemoglobin density in red blood cells rather than whole blood. Because specific volume is the inverse of density, the hemoglobin specific volume can be calculated by MCHC. However, hemoglobin specific volume and HSW are different in that the former emphasizes the specific volume, while the latter focuses more on the fluctuation of the specific volume. So the RBC volumetric fluctuation parameter needs to be taken into account. Interestingly, the RDW ^20^ was used to describe the variability or fluctuation of red blood cell volume. That is, the HSW can be calculated with RDW and HSV. In summary, the rude calculation strategy of HSW is finally formulated, and HSW is equal to RDW divided by MCHC. While the calculation strategy of HSW was developed, more details are needed to justify it.

So, the calculated detail of HSW is shown in **Fig. 1**. First, RDW equals erythrocyte volume’s standard deviation (EV-SD) by the mean corpuscular volume (MCV) ^21^. Second, the MCHC ^22^ equals mean cell hemoglobin (MCH) divided by the mean corpuscular volume (MCV). And HSW equals RDW divided by MCHC. With the process of **Fig. 1**, the HSW is finally equal to EV-SD divided by MCH. From the formula, HSW can characterize fluctuations in red blood cell volume per gram of hemoglobin.

**Fig.1.**
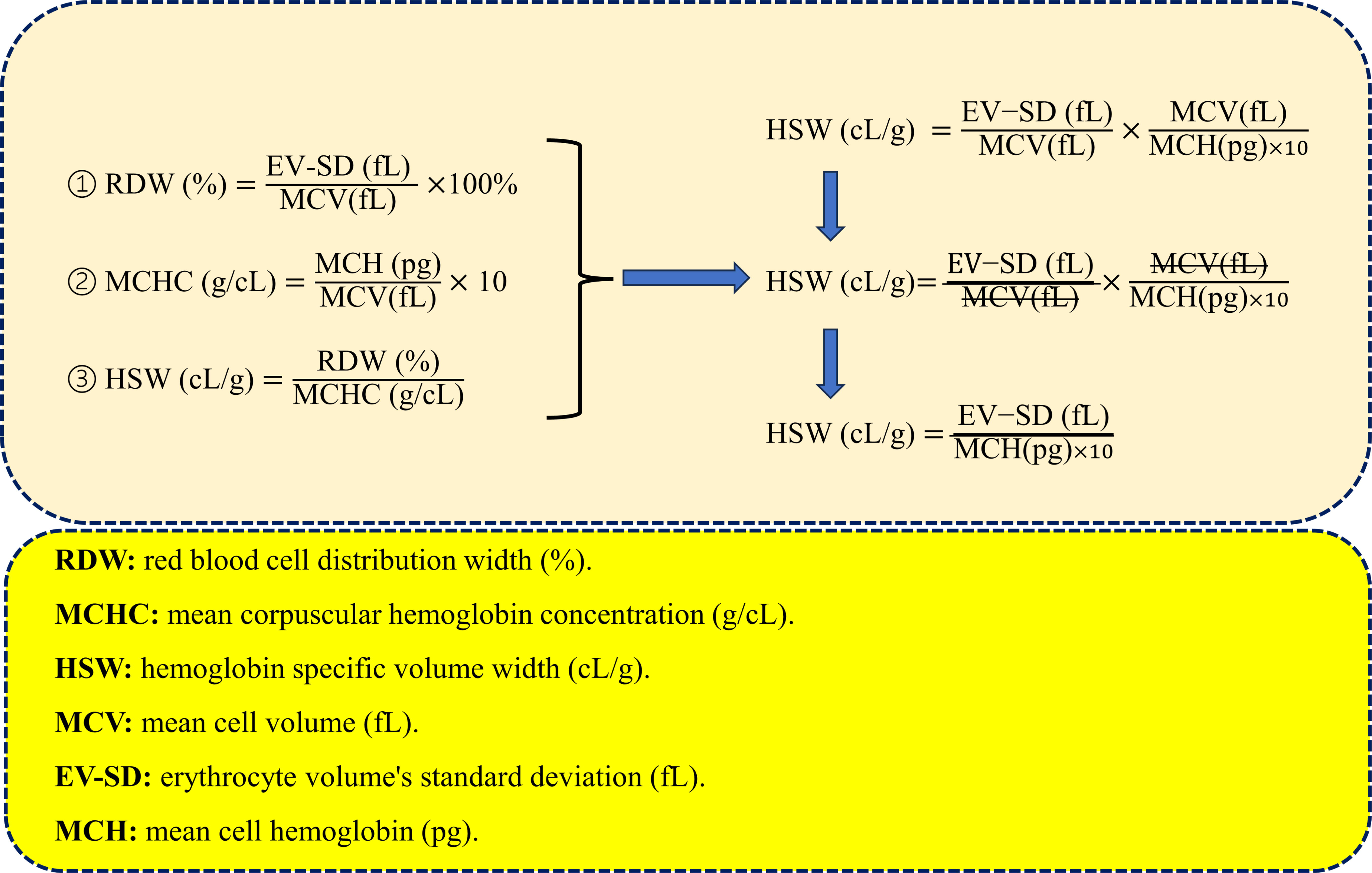
The calculated detail of HSW.

### 2.2 Participants

National Health and Nutrition Examination Survey (NHANES) 1999–2020 offer the primary data. NHANES, as a periodic health-related survey in America, currently contained 111,797 patients and was open to the public free of charge (www.cdc.gov/nchs/nhanes/irba98.htm.). All the research participants provided written informed consent.

In NHANES, the inclusion criteria included age >18 years, HSW presence, answering the CVD questionaries, and completing lab tests. The following were excluded: age <= 18 years (*n*=45,229), without a questionnaire answering whether there was a history of CVD (*n*=2,267), individuals without primary circle weights (*n* = 3,308), the missing HSW (*n*=3,099). Therefore, a total of 57,894 participants were included in the following process. Of the 57,894 participants, 6,651 CVD patients were specifically filtered for prognosis analysis. And the excluded 1,028 CVD patients without follow-up time. Finally, 57,894 participants were included for prevalence analysis, and 5,623 CVD patients were included in the prognosis process (**Fig. 2**).

**Fig. 2.**
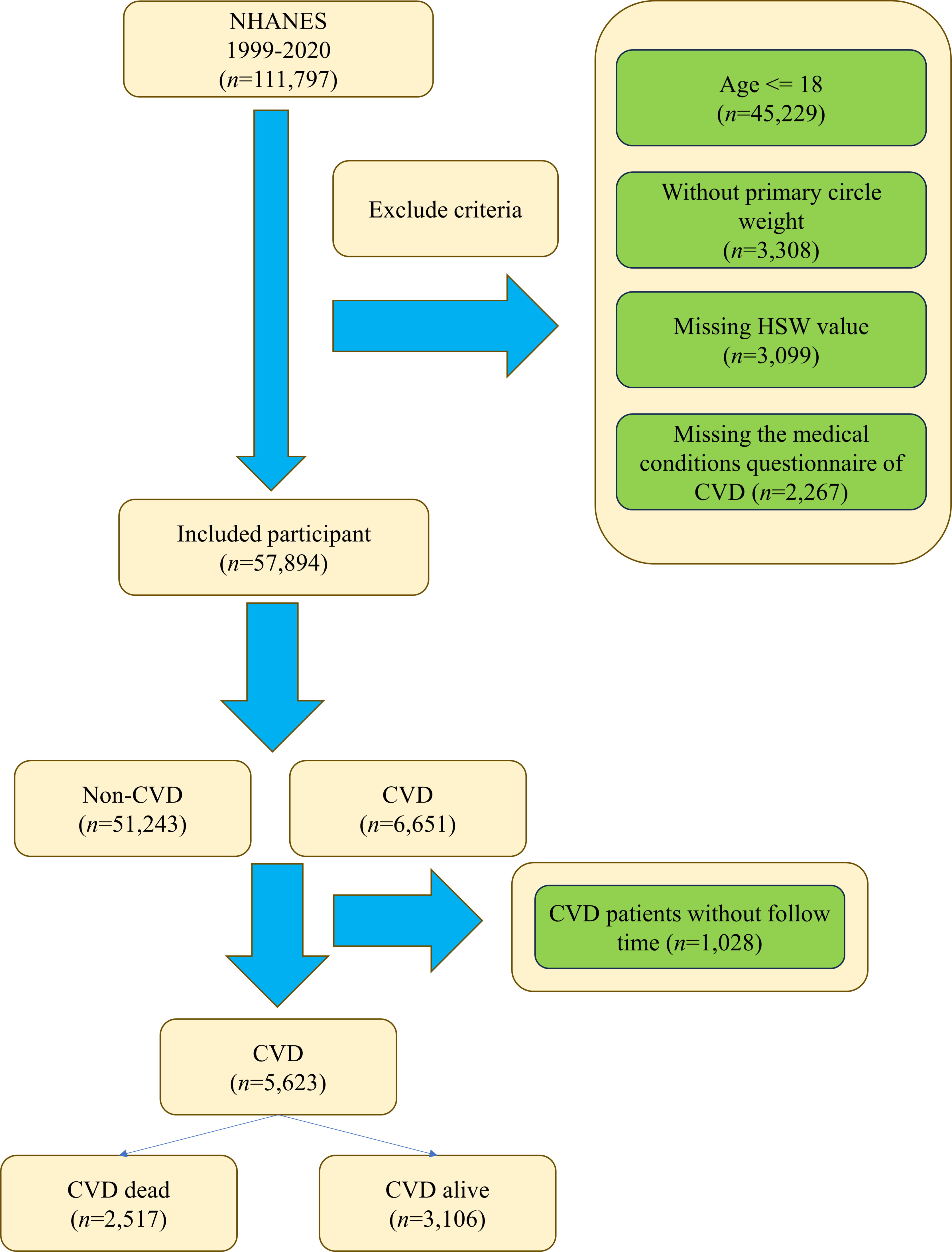
The flow chart selection from the NHANES 1999-2020.

### 2.3 The definition of CVD

The medical conditions questionnaire (MCQ) mainly obtained the relevant diagnoses (https://www.n.cdc.gov/nchs/nhanes/default.aspx). CVD in these studies was defined as any of the following five diseases: coronary heart disease, congestive heart failure, heart attack, stroke, and angina. The questionnaire codes in the MCQs are different for different diseases. For example, the MCQ codes for coronary heart disease were MCD180c, MCQ160c, and MCQ180c. And congestive heart failure MCQ codes were MCD180b, MCQ160b, and MCQ180b. Next, the heart attack MCQ codes were MCD180e, MCD160e, and MCQ180e. The MCQ codes of stroke were MCD180f, MCQ160f, and MCQ180f. Finally, the codes of angina were MCD180d, MCQ160d, and MCQ180d.

### 2.4 Covariates

Several covariates were filtered based on the following criteria, i) demographic variables; ii) characteristics affecting CVD reported in previous studies; and iii) other variables derived from the clinical experience.

Previous research emphasized that several diseases are associated with CVD, including chronic kidney disease (CKD) ^23^, chronic obstructive pulmonary disease (COPD) ^24–26^, hypertension ^27^, and diabetes mellitus (DM) ^28^. CKD-EPI ^29^ formulas were applied to access the estimated glomerular filtration rate (eGFR) for CKD diagnosis. Based on guideline ^30^, the eGFR < 90 mL/min was classified into the CKD group, else the non-CKD group. COPD was diagnosed with four criteria, FEV1/FVC < 0.7 in post-bronchodilator, the MCQ coded of MCQ160g and MCQ160p that ever told you had emphysema, age above 40 with a smoke history or chronic respiratory diseottom, and the history of COPD drugs (*e,g.* selective phosphodiesterase-4 inhibitors, mast cell stabilizers, and leukotriene modifiers). The diagnosis of hypertension with systolic blood pressure > 140 mmHg or diastolic > 90 mmHg. The diagnosis of DM includes five criteria, including glycohemoglobin (%) >= 6.5, fasting glucose (mmol/L) >= 7.0, random blood glucose (mmol/L) >= 11.1, two-hour oral glucose tolerance test blood glucose (mmol/L) >= 11.1, and history of drug (*e,g.* diabetes medication and insulin). Impaired fasting glycaemia and impaired glucose tolerance were divided into the preDM group.

In addition to the above covariates, other lab tests were tallied, including albumin (ALB, g/dL), alanine transaminase (ALT, U/L), basophils percentage (BaP, %), blood urea nitrogen (BUN, mmol/L), total calcium (Ca, mg/dL), chloride (Cl, mmol/L), eosinophils number count (Eo, 1000 cells/μL), eosinophils percentage (EoP, %), iron (Fe, ug/dL), gammaglutamyl transferase (GGT, U/L), globulin (GLB, g/dL), glucose (Glu, mg/dL), bicarbonate (HCO3, mmol/L), hematocrit (Hem, %), lymphocyte percentage (LymP, %), MCHC (g/cL), MCV (fL), monocyte number (Mon, 1000 cells/μL), monocyte percentage (MonP, %), mean platelet volume (MPV, fL), sodium (Na, mmol/L), platelet count count (Plt, 1000 cells/μL), RBC count (RBC, million cells/μL), RDW (%), segmented neutrophils percentage (SegneP, %), triglycerides (TG, mg/dL), segmented neutrophils number percentage (SeneP, %), total cholesterol (TC, mg/dL), uric acid (UA, mg/dL), total protein (TP, g/dL), and white blood cell count (WBC, 1000 cells/μL).

### 2.5 The all-caused mortality for CVD patients

For NHANES, National Death Index (https://www.cdc.gov/nchs/ndi/index.htm) was linked with NHANES and directly provided the mortality-related data. As previously reported ^31^, CVD all-cause mortality information was derived and matched with unique individuals’ IDs. Followup time was defined as the period of blood draws time to death or December 31, 2019.

### 2.6 Statistical analyses

All analysis processes were finished with R software (version 4.2.2). In cleaning clinical data, the R packages *nhanesR* (version 0.9.4.1) were served for NHANES.

In the analysis of variance, if the continuous variables were distributed with Gaussian, the Student’s t-test process, and else Mann–Whitney U. A chi-square test was performed to factor data (e.g., Sex and ethnicity). The continuous variables were presented as means ± SDs and proportions for factor variables.

Logistic regression was developed to explore the odds ratio (OR) in the non-CVD and CVD groups. Cox regression was applied to the HR and 95% confidence intervals (CIs) of HSW for CVD all-cause mortality. Time-to-death among groups is presented with Kaplan–Meier (KM) curves. Importantly, to optimize the cut-off for HSW in CVD patient mortality, restricted cubic spline (RCS) ^32^ was applied in this work. The RCS model (with 3-8 knots) examined the optimal HSW value association with CVD mortality. Only a *P*-value <0.05 was considered statistically significant.

## 3. Result

### 3.1 General characteristics

Among 57,894 participants were included, 30,052 (51.91%) females and 27,842 (48.09%) males, with an average age of 49.92, and 18,695 (32.29%) were over 60 years old. All the participants were classified into non-CVD (*n*=51,243) or CVD (*n*=6,651) individuals, which could represent 210,170,251 Americans (**Table 1**) according to the calculated weights. The HSW level in CVD groups (4.05±0.01) is higher than the non-CVD group (3.85±0.00). Among the 43 characteristics, only two had no significance, including Lym and AST.

**Table 1.** General characteristics for non-CVD and CVD in the NHANES.

| Variables | non-CVD (n=51,243) | CVD (n=6,651) | P-value |
| --- | --- | --- | --- |
| HSW, cL/g | 3.85±0.00 | 4.05±0.01 | < 0.0001 |
| Sex, (Male), % | 24084(47.44) | 3758(53.40) | < 0.0001 |
| Age, year | 45.58±0.16 | 64.82±0.25 | < 0.0001 |
| Ethnicity |  |  | < 0.0001 |
| Mexican American, % | 9032(8.62) | 728(4.12) |  |
| Non-Hispanic Black, % | 10834(10.80) | 1421(10.93) |  |
| Non-Hispanic White, % | 21353(67.24) | 3635(75.24) |  |
| Other Hispanic, % | 4534(6.06) | 427(3.60) |  |
| Other Race, % | 5490(7.28) | 440(6.11) |  |
| CKD, % | 7714(12.18) | 3007(40.95) | < 0.0001 |
| COPD, % | 1373( 3.07) | 677(12.83) | < 0.0001 |
| Diabetes |  |  | < 0.0001 |
| Health, % | 38566(82.06) | 3437(54.92) |  |
| preDM, % | 3519(6.78) | 514(8.74) |  |
| DM, % | 7720(11.16) | 2691(36.34) |  |
| Hypertension, % | 18913(32.89) | 5139(74.08) | < 0.0001 |
| WBC, 1000 cells/μL | 7.27±0.02 | 7.55±0.05 | < 0.0001 |
| LymP, % | 30.25±0.07 | 27.53±0.17 | < 0.0001 |
| MonP, % | 7.94±0.02 | 8.49±0.04 | < 0.0001 |
| SegneP, % | 58.39±0.08 | 60.20±0.18 | < 0.0001 |
| EoP, % | 2.77±0.01 | 3.10±0.03 | < 0.0001 |
| BaP, % | 0.72±0.00 | 0.75±0.01 | < 0.001 |
| Lym, 1000 cells/μL | 2.15±0.01 | 2.09±0.04 | 0.11 |
| Mon, 1000 cells/μL | 0.56±0.00 | 0.62±0.00 | < 0.0001 |
| SeneP, % | 4.32±0.02 | 4.56±0.03 | < 0.0001 |
| Eo, 1000 cells/μL | 0.20±0.00 | 0.23±0.00 | < 0.0001 |
| Ba, 1000 cells/μL | 0.05±0.00 | 0.05±0.00 | < 0.0001 |
| RBC, million cells/μL | 4.72±0.01 | 4.58±0.01 | < 0.0001 |
| Hg, g/dl | 14.31±0.02 | 14.00±0.03 | < 0.0001 |
| Hem, % | 42.14±0.05 | 41.44±0.09 | < 0.0001 |
| MCV, fL | 89.55±0.07 | 90.70±0.10 | < 0.0001 |
| MCH, pg | 30.40±0.03 | 30.65±0.05 | < 0.0001 |
| MCHC, g/cL | 3.39±0.00 | 3.38±0.00 | < 0.0001 |
| RDW, % | 13.05±0.01 | 13.65±0.02 | < 0.0001 |
| Plt, 1000 cells/μL | 254.64±0.62 | 235.92±1.36 | < 0.0001 |
| MPV, fL | 8.18±0.01 | 8.24±0.02 | 0.01 |
| ALB, g/dL | 4.27±0.00 | 4.12±0.01 | < 0.0001 |
| ALT, U/L | 25.31±0.12 | 23.65±0.32 | < 0.0001 |
| AST, U/L | 24.77±0.09 | 25.00±0.23 | 0.35 |
| BUN, mmol/L | 13.35±0.05 | 17.45±0.14 | < 0.0001 |
| Ca, mg/dL | 9.41±0.01 | 9.39±0.01 | 0.02 |
| TC, mg/dL | 196.57±0.38 | 184.27±0.93 | < 0.0001 |
| HCO<sub>3</sub>, mmol/L | 24.85±0.04 | 25.12±0.06 | < 0.0001 |
| GGT, U/L | 28.17±0.23 | 34.20±0.65 | < 0.0001 |
| Glu, mg/dL | 97.10±0.20 | 112.97±0.68 | < 0.0001 |
| Fe, ug/dL | 88.00±0.27 | 81.61±0.58 | < 0.0001 |
| TP, g/dL | 7.16±0.01 | 7.06±0.01 | < 0.0001 |
| TG, mg/dL | 147.88±1.03 | 167.43±2.02 | < 0.0001 |
| UA, mg/dL | 5.34±0.01 | 5.89±0.03 | < 0.0001 |
| Na, mmol/L | 139.35±0.06 | 139.46±0.08 | 0.03 |
| Cl, mmol/L | 103.08±0.06 | 102.58±0.08 | < 0.0001 |
| GLB, g/dL | 2.89±0.01 | 2.94±0.01 | < 0.0001 |

HSW’s quartiles (HSWQ) were determined. The HSW were classified into Q1 [1.88,3.64] cL/g, Q2 (3.64,3.85] cL/g, Q3 (3.85,4.11] cL/g, and Q4 (4.11,11.74] cL/g. Details of the HSW quartiles were shown in **Table 2**. CVD prevalence in HSW quartiles was 4.95%, 7.11%, 10.51%, and 16.14%, respectively. The percentage of CVD increases with increasing HSW levels. And increasing HSW levels accompany decreasing males percentage and increased age.

**Table 2.** General characteristics for quartiles of HSW.

| **Variables** | **Q1** | **Q2** | **Q3** | **Q4** | **P-value** |
| --- | --- | --- | --- | --- | --- |
| **HSW range, cL/g** | **(n=14,462)** | **(n=14,369)** | **(n=14,579)** | **(n=14,484)** | **e** |
|  | **[1.88,3.64]** | **(3.64,3.85]** | **(3.85,4.11]** | **(4.11,11.74]** |  |
| HSW, cL/g | 3.49±0.00 | 3.74±0.00 | 3.96±0.00 | 4.53±0.01 | < 0.0001 |
| CVD, % | 909(4.95) | 1303(7.11) | 1805(10.51) | 2634(16.14) | < 0.0001 |
| Sex, (Male), % | 7321(50.98) | 7419(51.82) | 7263(48.68) | 5839(37.06) | < 0.0001 |
| Age, year | 42.72±0.25 | 46.34±0.24 | 50.08±0.28 | 52.46±0.29 | < 0.0001 |
| Ethnicity |  |  |  |  | < 0.0001 |
| Mexican American, % | 3006(8.17) | 2662(8.40) | 2311(8.52) | 1781(7.64) |  |
| Non-Hispanic Black, % | 1348( 4.49) | 2237( 8.05) | 3227(11.92) | 5443(23.25) |  |
| Non-Hispanic White, % | 7855(75.81) | 6787(71.32) | 5881(65.33) | 4465(54.14) |  |
| Other Hispanic, % | 1094(5.10) | 1190(5.58) | 1456(6.58) | 1221(6.45) |  |
| Other Race, % | 1159(6.43) | 1493(6.65) | 1704(7.65) | 1574(8.51) |  |
| CKD, % | 1645( 9.03) | 2226(12.67) | 2855(16.62) | 3995(24.49) | < 0.0001 |
| COPD, % | 418(3.06) | 438(3.33) | 516(4.13) | 678(6.25) | < 0.0001 |
| Diabetes state |  |  |  |  | < 0.0001 |
| Health, % | 11480(86.14) | 10866(82.24) | 10348(76.37) | 9309(69.56) |  |
| preDM, % | 832(5.51) | 1001(6.66) | 1168(8.47) | 1032(7.78) |  |
| DM, % | 1654( 8.35) | 2094(11.10) | 2780(15.16) | 3883(22.66) |  |
| Hypertension, % | 4576(29.14) | 5427(33.72) | 6360(39.92) | 7689(48.23) |  |
| WBC, 1000 cells/μL | 7.14±0.03 | 7.20±0.03 | 7.39±0.04 | 7.57±0.04 | < 0.0001 |
| LymP, % | 30.26±0.10 | 30.33±0.10 | 29.94±0.11 | 29.23±0.15 | < 0.0001 |
| MonP, % | 7.85±0.03 | 7.99±0.03 | 8.05±0.03 | 8.13±0.03 | < 0.0001 |
| SegneP, % | 58.48±0.11 | 58.22±0.12 | 58.52±0.13 | 59.16±0.16 | < 0.0001 |
| EoP, % | 2.78±0.02 | 2.81±0.02 | 2.83±0.02 | 2.79±0.02 | < 0.0001 |
| BaP, % | 0.67±0.01 | 0.71±0.01 | 0.75±0.01 | 0.78±0.01 | 0.29 |
|  |  |  |  |  | < |
| Lym, 1000 cells/μL | 2.11±0.01 | 2.13±0.01 | 2.17±0.02 | 2.19±0.02 | 0.0001 |
| Mon, 1000 cells/μL | 0.55±0.00 | 0.56±0.00 | 0.58±0.00 | 0.59±0.00 | 0.001 |
|  |  |  |  |  | < |
| SeneP, % | 4.24±0.02 | 4.26±0.02 | 4.39±0.03 | 4.54±0.03 | 0.0001 |
|  |  |  |  |  | < |
| Eo, 1000 cells/μL | 0.20±0.00 | 0.20±0.00 | 0.20±0.00 | 0.21±0.00 | 0.0001 |
|  |  |  |  |  | < |
| Ba, 1000 cells/μL | 0.04±0.00 | 0.04±0.00 | 0.05±0.00 | 0.05±0.00 | 0.001 |
|  |  |  |  |  | < |
| RBC, million cells/μL | 4.67±0.01 | 4.72±0.01 | 4.73±0.01 | 4.69±0.01 | 0.0001 |
|  |  |  |  |  | < |
| Hg, g/dl | 14.69±0.03 | 14.53±0.02 | 14.29±0.02 | 13.27±0.03 | 0.0001 |
|  |  |  |  |  | < |
| Hem, % | 42.48±0.08 | 42.67±0.06 | 42.42±0.07 | 40.18±0.08 | 0.0001 |
|  |  |  |  |  | < |
| MCV, fL | 91.06±0.08 | 90.50±0.06 | 89.89±0.08 | 85.96±0.13 | 0.0001 |
|  |  |  |  |  | < |
| MCH, pg | 31.49±0.03 | 30.80±0.03 | 30.28±0.03 | 28.40±0.05 | 0.0001 |
|  |  |  |  |  | < |
| MCHC, g/cL | 3.46±0.00 | 3.40±0.00 | 3.37±0.00 | 3.30±0.00 | 0.0001 |
|  |  |  |  |  | < |
| RDW, % | 12.07±0.01 | 12.72±0.01 | 13.36±0.01 | 14.93±0.02 | 0.0001 |
|  |  |  |  |  | < |
| Plt, 1000 cells/μL | 254.85±0.87 | 250.45±0.83 | 247.93±0.86 | 259.63±1.05 | 0.0001 |
|  |  |  |  |  | < |
| MPV, fL | 8.06±0.02 | 8.17±0.01 | 8.25±0.01 | 8.33±0.02 | 0.0001 |
|  |  |  |  |  | < |
| ALB, g/dL | 4.36±0.01 | 4.30±0.00 | 4.22±0.01 | 4.08±0.01 | 0.0001 |
|  |  |  |  |  | < |
| ALT, U/L | 26.69±0.18 | 25.80±0.29 | 24.80±0.22 | 22.27±0.22 | 0.0001 |
|  |  |  |  |  | < |
| AST, U/L | 25.50±0.14 | 24.79±0.16 | 24.52±0.17 | 23.96±0.17 | 0.0001 |
|  |  |  |  |  | < |
| BUN, mmol/L | 12.90±0.06 | 13.55±0.06 | 14.17±0.09 | 14.69±0.09 | 0.0001 |
|  |  |  |  |  | < |
| Ca, mg/dL | 9.47±0.01 | 9.43±0.01 | 9.39±0.01 | 9.32±0.01 | 0.0001 |
|  |  |  |  |  | < |
| TC, mg/dL | 197.36±0.51 | 197.53±0.57 | 195.20±0.49 | 189.89±0.75 | 0.0001 |
|  |  |  |  |  | < |
| HCO3, mmol/L | 24.71±0.05 | 24.94±0.06 | 25.00±0.06 | 24.89±0.06 | 0.0001 |
|  |  |  |  |  | < |
| GGT, U/L | 28.32±0.35 | 27.73±0.32 | 29.51±0.57 | 29.67±0.43 | 0.0001 |
| Glu, mg/dL | 95.57±0.35 | 97.68±0.31 | 100.21±0.37 | 102.32±0.36 | <<br>0.001 |
| Fe, ug/dL | 96.54±0.44 | 91.26±0.47 | 85.92±0.44 | 69.35±0.49 | <<br>0.0001 |
| TP, g/dL | 7.20±0.01 | 7.16±0.01 | 7.12±0.01 | 7.12±0.01 | <<br>0.0001 |
| TG, mg/dL | 159.23±1.82 | 147.96±1.46 | 147.02±1.51 | 139.80±1.42 | <<br>0.0001 |
| UA, mg/dL | 5.36±0.02 | 5.40±0.01 | 5.41±0.02 | 5.42±0.02 | <<br>0.0001 |
| Na, mmol/L | 139.09±0.05 | 139.35±0.06 | 139.56±0.09 | 139.56±0.10 | 0.04 |
| Cl, mmol/L | 103.31±0.07 | 103.04±0.06 | 102.84±0.07 | 102.85±0.08 | <<br>0.0001 |
| GLB, g/dL | 2.83±0.01 | 2.87±0.01 | 2.90±0.01 | 3.04±0.01 | <<br>0.0001 |

### 3.2 Logistic regression for HSW and CVD

The univariate and adjusted Logistic details are shown in **Table S1.** The result of HSW, HSWQ, RDW, and MCHC were shown in **Table 3**. Three models were utilized in the adjustment. The ORs value of HSW in the three multivariant adjusted models were still significant, including 1.73(95% CI 1.62,1.86), 1.52(95% CI 1.40,1.65), and 1.46(95% CI 1.33,1.60). Taking Q1 as a reference and adjusting with multivariable (**Table 3**), Q4 was still significant and had the highest ORs, 1.54(95% CI 1.36,1.73). In summary, higher HSW might be a risk of CVD and might promote the prevalence of CVD.

**Table 3.**
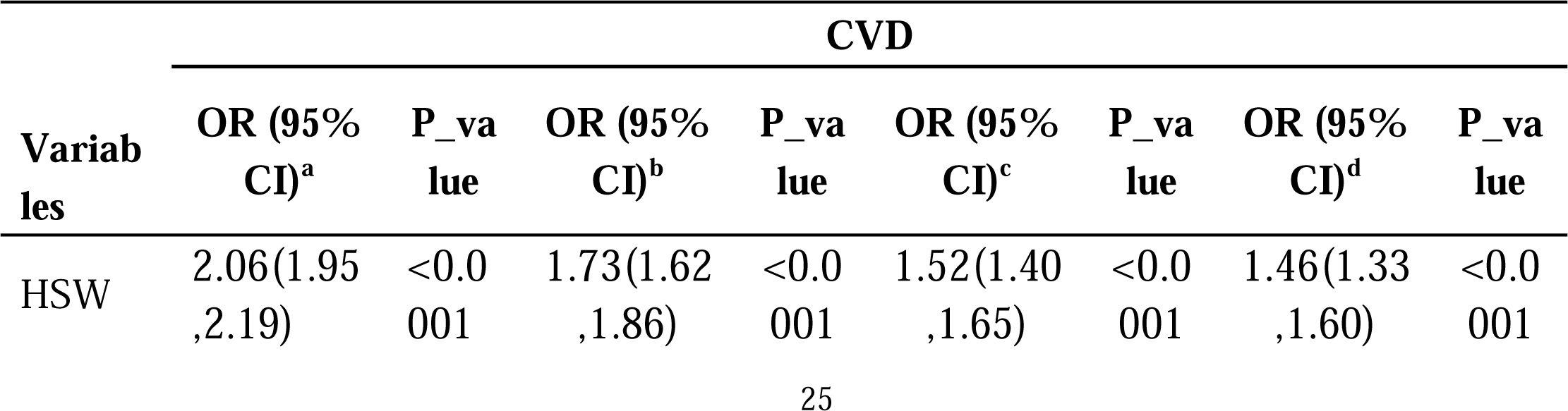
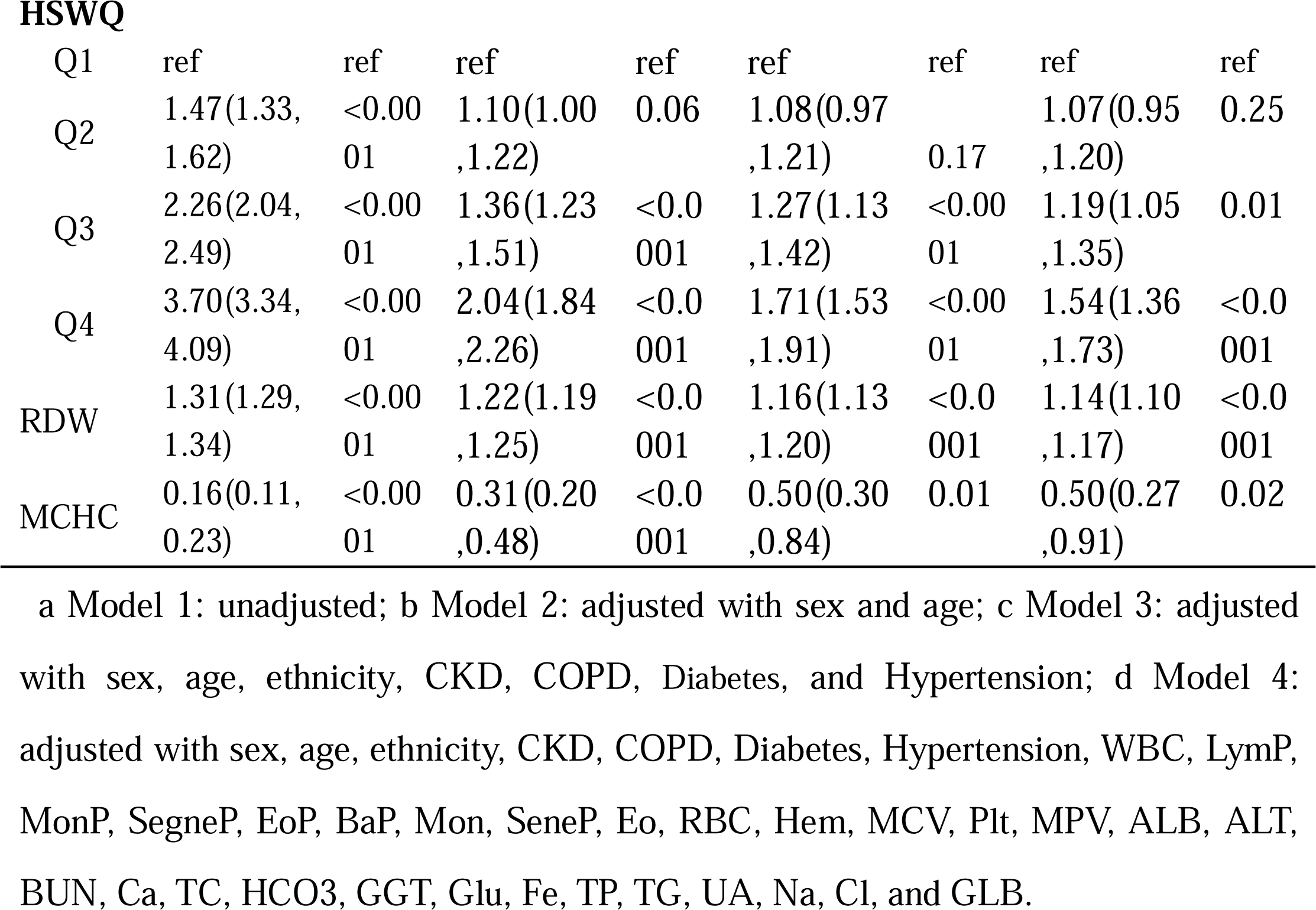
The univariate and adjusted Logistic congression model among HSW, HSWQ, RDW, and MCHC.

More importantly, The OR of RDW was consistently smaller than HSW’s, both univariate or adjusted. For example, the adjusted ORs with Model 4 in HSW and RDW were 1.46(95% CI 1.33,1.60) and 1.14(95% CI 1.10,1.17). In conclusion, higher HSW is a risk for the evolution of CVD with a much higher predictive value than RDW.

### 3.3 Logistic regression for HSW and CVD with subgroup

#### 3.3.1 Correlation of HSW and CVD with age and sex level

As shown in **Tables S2** and **S3,** the prevalence of CVD is significantly related to the stratification of age and sex. When adjusted with multivariate, HSW was still significantly correlated with CVD in females (OR: 1.49; 95% CI 1.32, 1.68) and males (OR: 1.37; 95% CI 1.18, 1.60). The OR values of females were higher than that of males, indicating females might be more at risk for CVD with an increasing HSW level. In the age with multivariable adjustment, the association between HSW and CVD in patients <= 60 years old (OR: 1.57; 95% CI 1.35–1.82) was higher than that in those > 60 years old (OR: 1.35; 95% CI 1.18–1.53). Both Sex and age in HSW, with Q1 as a reference and adjusted with multivariate, Q4 was still significantly associated with CVD.

#### 3.3.2 Correlation of HSW and CVD with various diabetes states

As shown in **Table S4,** three diabetes state was classified in this study, including healthy individuals, preDM, and DM patients. Among the three states, the HSW OR of DM was the highest and still significant after the multivariate adjustment (OR: 1.54; 95% CI 1.30–1.84). Processing Q1 as reference, Q4 was still associated with CVD in DM (OR: 1.77; 95% CI 1.36–2.29). At high levels of HSW, patients with DM will be more likely to develop CVD than healthy and prediabetic patients.

### 3.4 The Cox regression

Among the 6,651 CVD patients, 1,028 were excluded because of the miss the following information. 2,517 CVD patients died during the follow-up time. Follow-up time ranged from 1 to 249 months, and the median and interquartile spacing of follow-up times were 79 (40, 130). The general information on dead and alive CVD was shown in **Table 4**. The HSW level in dead CVD groups (4.06±0.01) is higher than in the alive CVD group (4.00±0.01). And the dead CVD group had a higher age, but the sex composition was insignificant.

**Table 4.** The general information on dead and alive CVD patients.

| Variables | CVD patients alived (*n* = 3,106) | CVD patients dead (*n* = 2,517) | P-value |
| --- | --- | --- | --- |
| HSW, cL/g | 4.00±0.01 | 4.06±0.01 | <0.001 |
| Sex, (Male), % | 1678(54.41) | 1488(51.69) | 0.15 |
| Age, year | 60.44±0.31 | 71.50±0.31 | <0.0001 |
| Ethnicity |  |  | <0.0001 |
| Mexican American, % | 394(5.08) | 262(2.87) |  |
| Non-Hispanic Black, % | 716(11.83) | 437( 9.55) |  |
| Non-Hispanic White, % | 1488(71.70) | 1657(81.51) |  |
| Other Hispanic, % | 256(4.12) | 87(2.34) |  |
| Other Race, % | 252(7.27) | 74(3.73) |  |
|  |  |  | < |
| CKD, % | 1048(29.55) | 1521(59.60) | 0.0001 |
|  |  |  | < |
| COPD, % | 322(11.13) | 355(15.46) | 0.001 |
|  |  |  | < |
| Diabetes state |  |  | 0.0001 |
| Health, % | 1175(32.73) | 1070(40.16) |  |
| preDM, % | 1673(58.53) | 1245(50.95) |  |
| DM, % | 251(8.74) | 201(8.88) |  |
|  |  |  | < |
| Hypertension, % | 2350(71.30) | 2019(79.10) | 0.0001 |
| WBC, 1000 cells/μL | 7.47±0.08 | 7.65±0.06 | 0.09 |
|  |  |  | < |
| LymP, % | 28.71±0.22 | 25.80±0.21 | 0.0001 |
| MonP, % | 8.39±0.06 | 8.59±0.06 | 0.01 |
|  |  |  | < |
| SegneP, % | 59.12±0.23 | 61.84±0.23 | 0.0001 |
| EoP, % | 3.10±0.05 | 3.13±0.05 | 0.75 |
|  |  |  | < |
| BaP, % | 0.76±0.01 | 0.69±0.01 | 0.0001 |
| Lym, 1000 cells/μL | 2.15±0.06 | 1.99±0.04 | 0.03 |
|  |  |  | < |
| Mon, 1000 cells/μL | 0.60±0.01 | 0.64±0.01 | 0.0001 |
|  |  |  | < |
| SeneP, % | 4.43±0.04 | 4.74±0.04 | 0.0001 |
| Eo, 1000 cells/μL | 0.23±0.00 | 0.23±0.00 | 0.3 |
|  |  |  | < |
| Ba, 1000 cells/μL | 0.05±0.00 | 0.05±0.00 | 0.0001 |
|  |  |  | < |
| RBC, million cells/μL | 4.65±0.01 | 4.46±0.02 | 0.0001 |
|  |  |  | < |
| Hg, g/dl | 14.18±0.04 | 13.76±0.05 | 0.0001 |
|  |  |  | < |
| Hem, % | 41.88±0.12 | 40.75±0.14 | 0.0001 |
|  |  |  | < |
| MCV, fL | 90.19±0.12 | 91.72±0.15 | 0.0001 |
|  |  |  | < |
| MCH, pg | 30.53±0.06 | 30.99±0.07 | 0.0001 |
| MCHC, g/cL | 3.38±0.00 | 3.38±0.00 | 0.23 |
|  |  |  | < |
| RDW, % | 13.50±0.03 | 13.69±0.04 | 0.001 |
| Plt, 1000 cells/μL | 236.48±1.78 | 237.77±2.04 | 0.6 |
| MPV, fL | 8.26±0.03 | 8.17±0.03 | 0.02 |
|  |  |  | < |
| ALB, g/dL | 4.17±0.01 | 4.10±0.01 | 0.0001 |
| ALT, U/L | 25.10±0.43 | 22.27±0.68 | 0.001 |
| AST, U/L | 25.36±0.31 | 25.50±0.40 | 0.78 |
|  |  |  | < |
| BUN, mmol/L | 15.60±0.16 | 19.96±0.25 | 0.0001 |
| Ca, mg/dL | 9.40±0.01 | 9.42±0.01 | 0.35 |
| TC, mg/dL | 184.04±1.29 | 188.08±1.32 | 0.02 |
| HCO<sub>3</sub>, mmol/L | 25.07±0.08 | 25.03±0.09 | 0.65 |
| GGT, U/L | 32.46±0.78 | 36.36±1.26 | 0.01 |
|  |  |  | < |
| Glu, mg/dL | 110.62±0.89 | 116.16±1.26 | 0.001 |
|  |  |  | < |
| Fe, ug/dL | 83.39±0.87 | 78.91±0.81 | 0.001 |
|  |  |  | < |
| TP, g/dL | 7.04±0.01 | 7.12±0.02 | 0.001 |
| TG, mg/dL | 169.74±2.65 | 166.63±3.10 | 0.41 |
|  |  |  | < |
| UA, mg/dL | 5.75±0.04 | 6.19±0.05 | 0.0001 |
| Na, mmol/L | 139.47±0.11 | 139.12±0.10 | 0.01 |
|  |  |  | < |
| Cl, mmol/L | 102.98±0.10 | 102.42±0.11 | 0.0001 |
|  |  |  | < |
| GLB, g/dL | 2.88±0.01 | 3.01±0.02 | 0.0001 |

Other information among the quartiles of HSW among these CVD patients is shown in **Table 5**. The mortality rates of the Q1, Q2, Q3, and Q4 were 35.80%, 41.77%, 35.28%, and 43.68% (*P*<0.001). Mortality fluctuates as the quantile of HSW increases. But HSW, the fourth percentile overall, predicts the highest mortality.

**Table 5.**
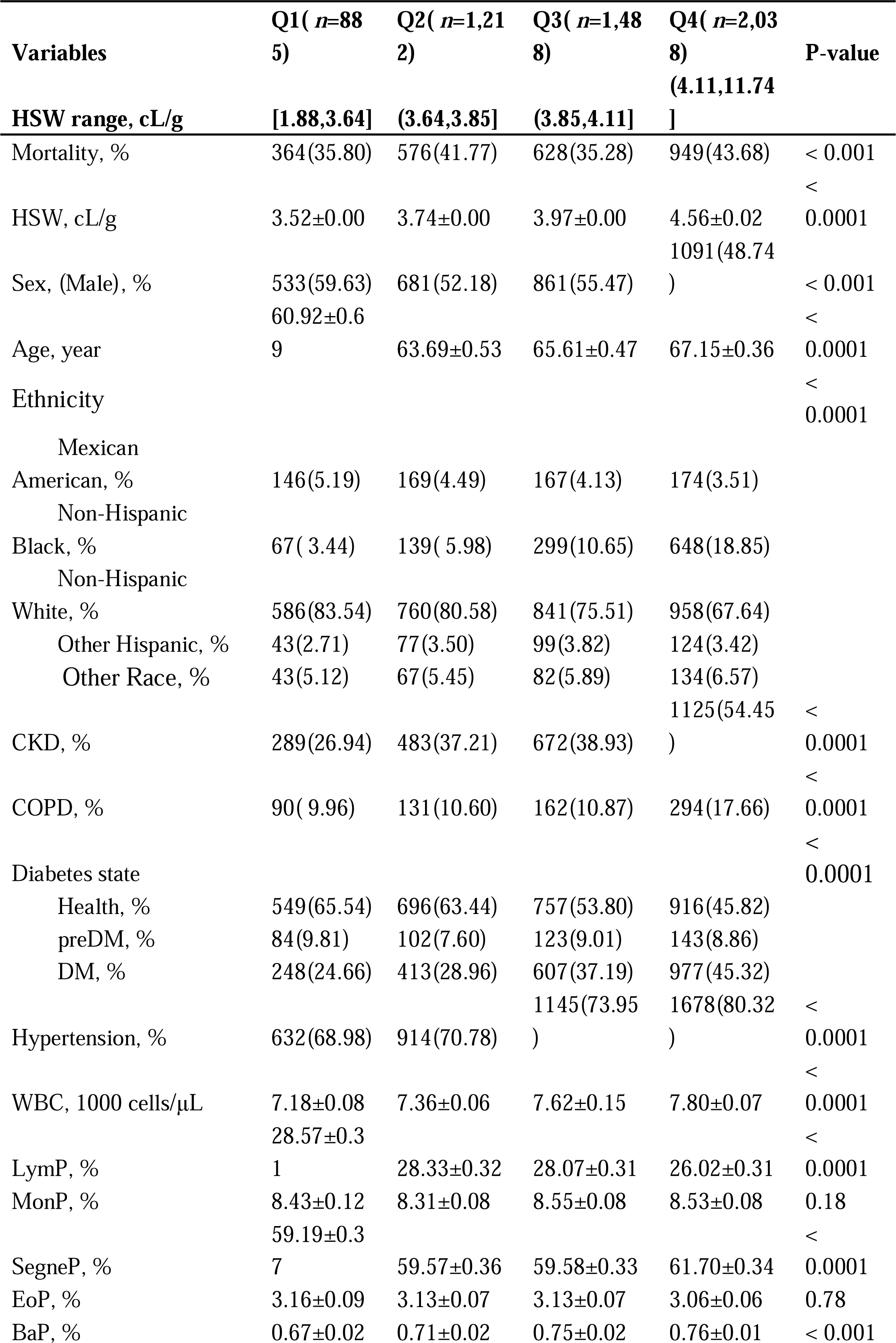
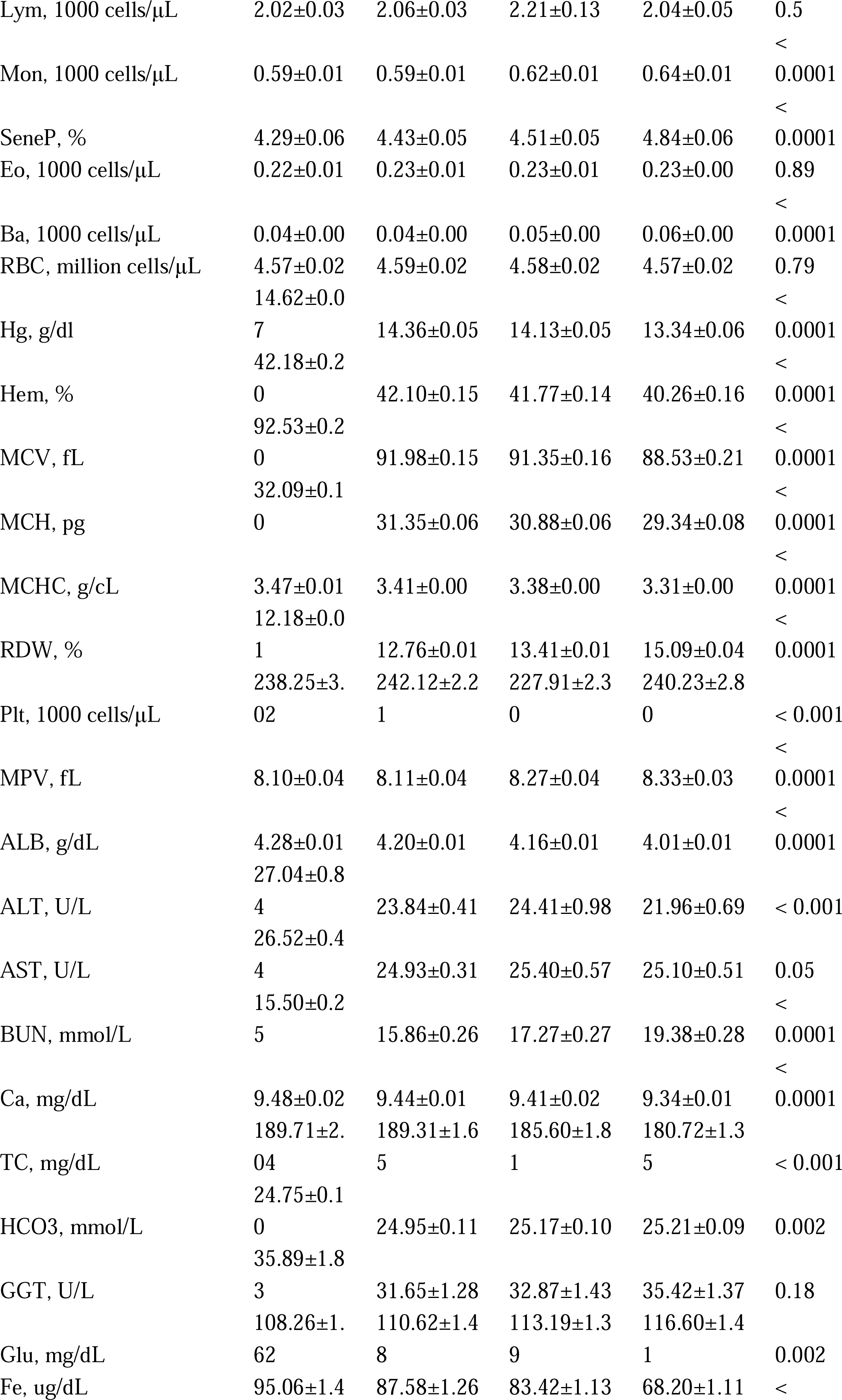
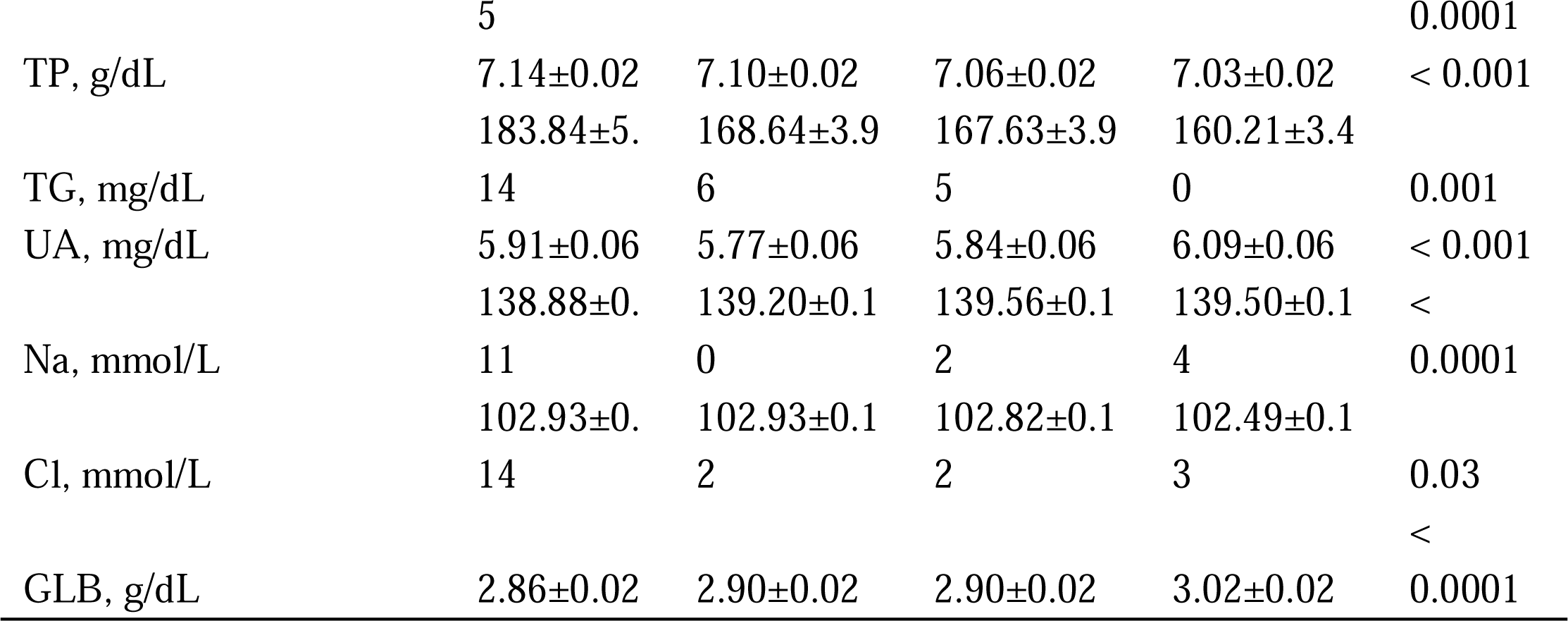
The general information of HSW quartiles among CVD patients.

The univariate and adjusted Cox regression results was shown in **Table S5.** The details of HSW, HSWQ, RDW, and MCHC were shown in **Table 6**. Adjusted with multivariable, the HR value of HSW was still significant, 1.70(95% CI 1.48,1.95). Using Q1 as a reference and adjusted with multivariable (**Table 6**), Q4 still had the significant and highest HRs, 1.88(95% CI 1.53, 2.31). The higher the HSW quartile was, the poor the CVD participants’ prognosis might be.

**Table 6.**
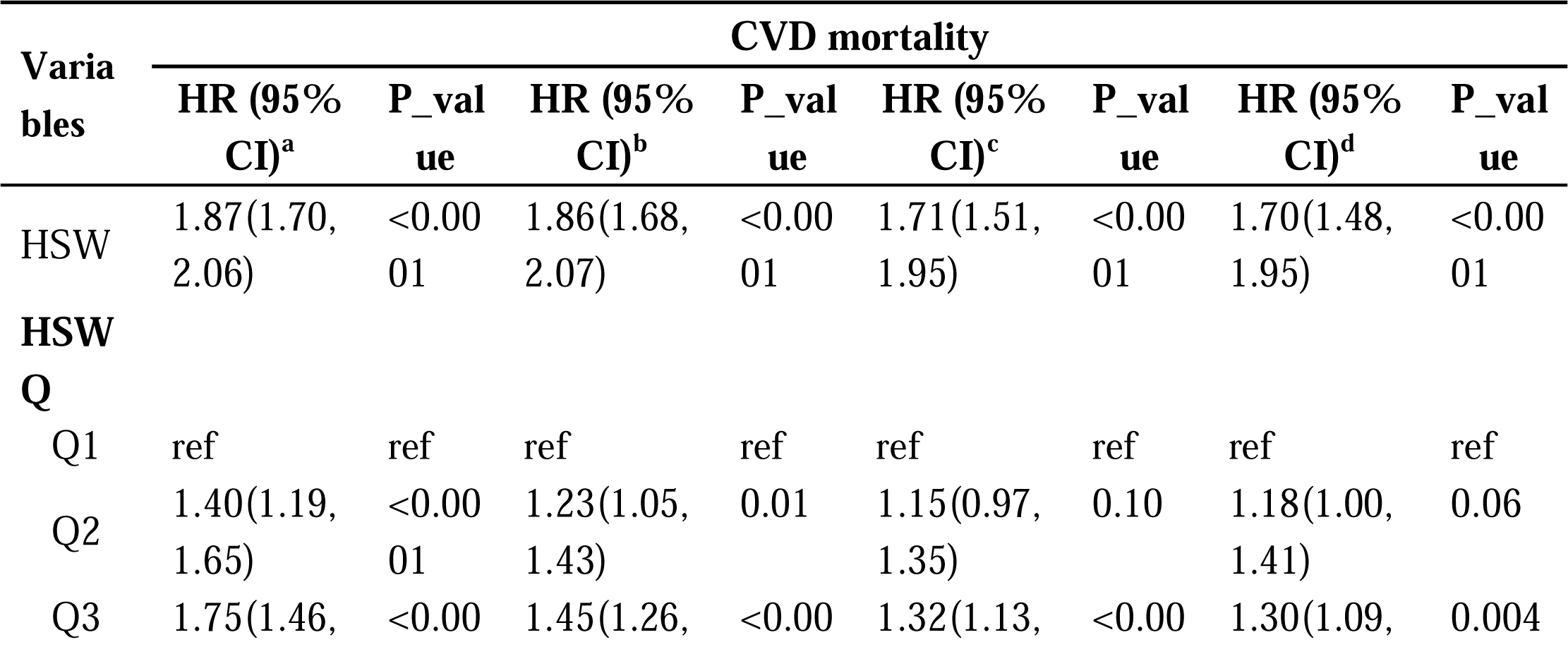
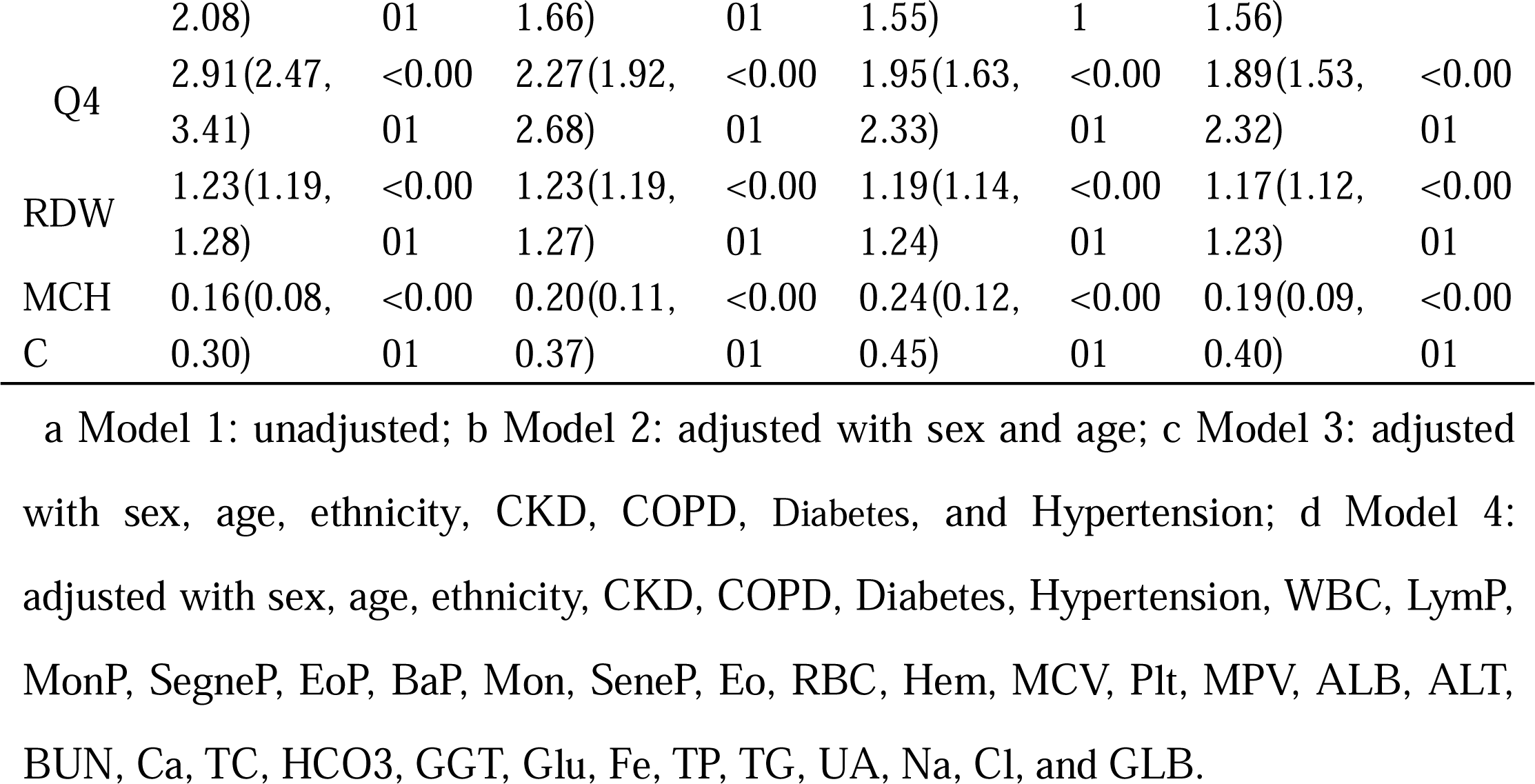
The univariate and adjusted Cox congression model among HSW, HSWQ, RDW, and MCHC.

More importantly, The HR of RDW was consistently smaller than HSW’s, both univariate or adjusted. For example, the adjusted HRs with the same multivariable in HSW and RDW were 1.70(95% CI 1.48,1.95) and 1.17(95% CI 1.12,1.23). In conclusion, higher HSW is an risk for the prognosis of CVD, accompanying a much higher predictive value than RDW.

### 3.5 RCS analysis and KM survival curve

In RCS analysis, according to the minimum fluctuation HR value range of 95% CI, the optimal value of HSW, MCHC, and RDW was determined (**Fig 3. A**). Both the RDW and HSW were poor prognoses of CVD. The higher HSW and RDW accompany the increasing HRs (**Fig 3. A**). Inversely, higher MCHC accompany decreasing HRs. And the optimal inflexion point of HSW, MCHC, and RDW were 3.96, 3.37, and 13.38, respectively.

**Fig 3.**
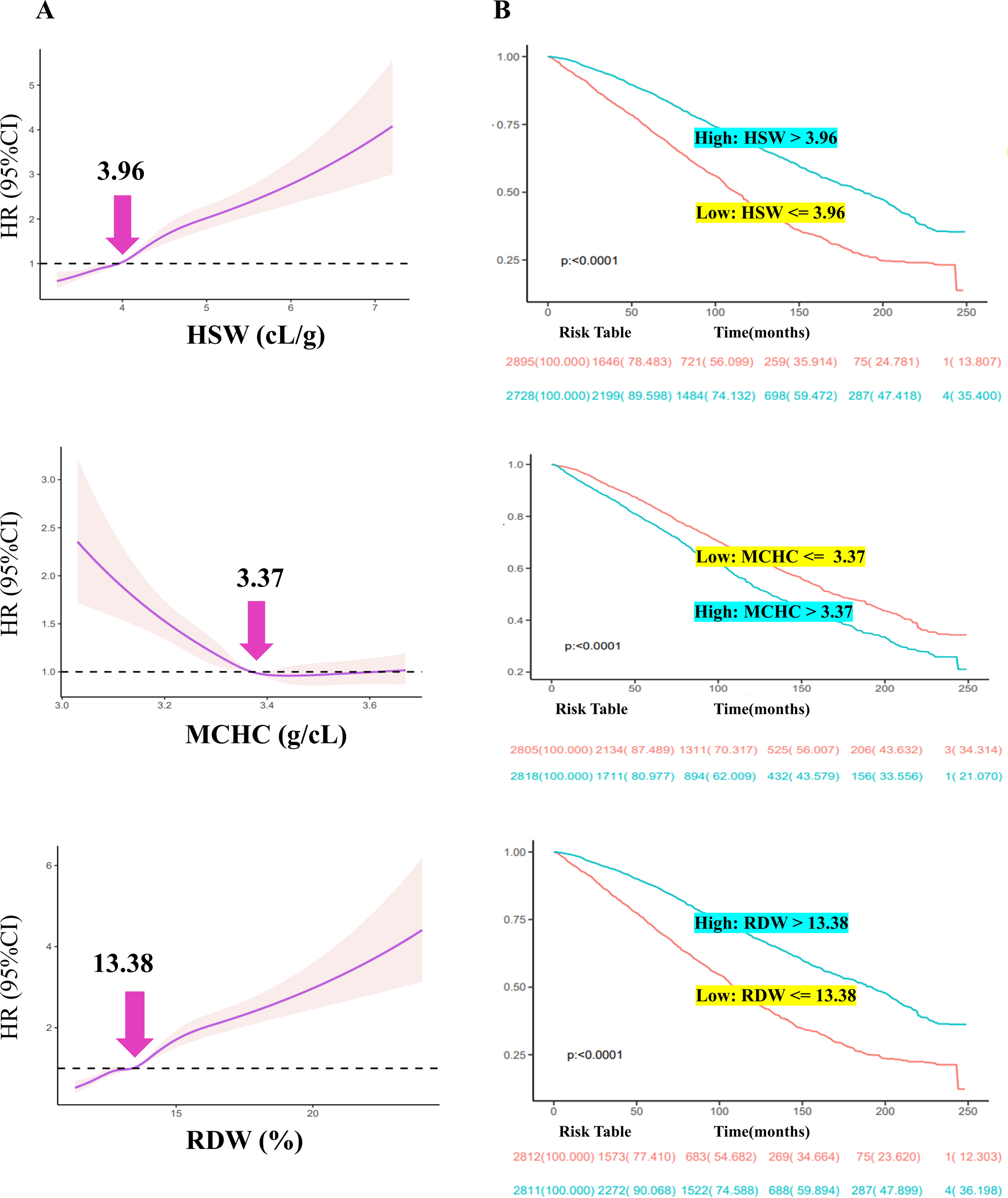
RCS analysis and KM survival curve. A The RCS for optimal cut-off HR value; B KM survival curve.

According to the optimal inflexion point, they were divided into the high-expression or low-expression groups. For example, an individual with an HSW >3.96 was classified into the high HSW group, else the low HSW group. Classified methods for RDW and MCHC were similar to HSW. CVD patients with HSW > 3.96, MCHC <3.37, and RDW > 13.38 have poor survival rates (**Fig 3. B**). The three factors stated an excellent predictive value in the survival analysis of CVD patients.

## 4. Discussion

This is the first article that focuses on the clinical value of HSW. It not only developed a calculation strategy for HSW but also verified the importance of HSW for the prevalence and long-term prognosis of CVD. The prevalence of CVD varied significantly among quartiles of HSW. Higher levels of HSW account for a higher prevalence of CVD. The risk of sex, age, and diabetes state stratification was different. Females had a higher risk of CVD than males when HSW increased. Individuals <=60 years old had a higher risk of CVD than those >60 when HSW grew. Compared with healthy and preDM, DM patients had a higher risk of CVD when higher HSW.

Furthermore, the prognosis of CVD changed significantly among quartiles of HSW and higher levels of HSW performance for a higher mortality rate of CVD. Moreover, an inverse L-curve with an inflexion point of 3.96 was observed between HR and CVD. And CVD patients with HSW > 3.96 indicated a higher mortality poor prognosis than HSW <= 3.96. These findings indicate that HSW can be used as a monitoring indicator of CVD incidence and prognosis.

Although HSW is calculated from RDW and MCHC, it has a much higher OR and HR value than both. For example, the adjusted ORs by Model 4 of HSW, RDW, and MCHC were 1.46, 1.14, and 0.50. For HRs, these were 1.70, 1.17, and 0.19. In a word, HSW is not only an independent predictive factor of the prevalence and prognosis of CVD, but the value is much higher than the RDW and MCHC.

The role of HSW in other diseases needs to be further investigated. For example, RDW ^33^ and MCHC ^34^ have already been recognized as diagnostic indicators of anaemia, and HSW is calculated from MCHC and RDW. MCHC mirrors the haemoglobin concentration and is related to an adverse poor prognosis in heart failure ^35^ (a subtype of CVD). Higher RDW ^36^ mirrors the potential deregulation of RBC balance, including weakened erythropoiesis and disorder RBC survival. Similarly, an increasing HSW might reflect a chaotic hematopoietic and oxygen-carrying. Because haemoglobin is an oxygen carrier, a rising HSW might mean an insufficient oxygen supply ^37^, which could trigger bad results, such as heart failure ^38,39^. Therefore, HSW might have a specific value in anaemia and hypoxic sensitivity diseases, and more studies are needed for in-depth analysis.

RDW has received an increasing focus on CVD over the last decade. Higher RDW might result in thrombotic disorders ^8^. A cross-sectional study with 495 individuals ^40^ emphasized that a combination of high RDW and thrombophilia abnormalities had a risk of cerebral vein thrombosis with 33.20 ORs. In a 15-year follow-up retrospective cohort with more than twenty thousand adult participants ^41^ without a heart attack, stroke, or heart failure, increasing RDW was related to long-term heart failure incidence with an HR value of 1.47. Following the percutaneous coronary intervention, RDW is conducive to predicting prognosis in CVD patients ^42^. The erythrocyte membrane enriches abundant cholesterol ^43^. Supplementing RBC within the atherosclerotic plaque will deteriorate plaque evolution ^44^, resulting in CVD events.

MCHC was widely used to diagnose and treat anaemia ^45^. Morteza *et, al*. ^35^ compared various RBC indices to predict a 12-month prognosis in heart failure. Among the above, the MCHC was the most sensitive index, and RDW was the most specific ^35^.

Another RDW-related novel index, RAR (the RDW-serum albumin ratio), has received increasing attention as an inflammation-related indicator in recent years. However, RAR was only utilized for the short-term prognosis in diseases (30 days ^46^, 90 days ^47,48^, 1 year ^49^). In CVD patients, HSW had over 70 months of long-term prognosis. Although RAR has some clinical value ^47,50^, they are combined with multiple tests. For example, the RDW ^47,50^ is derived from hematology, but serum albumin is from biochemistry, which may increase the patient’s financial burden. Unlike the above tests, both RDW and MCHC were included in hematology. In a word, HSW is more clinically actionable, applicable, and less financially burdensome for CVD patients.

This study has four strengths: innovation of HSW, large sample size, robust and credible results, and a combination of cross-sectional and prospective studies. Firstly, HSW is proposed by us for the first time, and the calculation strategy is formulated. Secondly, this study’s sample size and duration time (1999-2020) are large enough. NHANES applied a circle years weight to represent the American population. Thirdly, 36 covariates were included for adjustment to ensure the robustness and reliability of the results. More importantly, NHANES employed professional and trained personnel with standardized procedures to reduce deviation, such as strict physical examination and standard questionnaires. Finally, the combination of cross-sectional and prospective studies focuses on the incidence and the prognosis between HSW and CVD. The optimal cut-off value for HSW was determined in prognosis analysis, and its long-term prognostic value was analyzed.

Furthermore, two limitations should be noted. First, the causality could not be obtained, and the current result should be approached carefully. Further prospective work is indispensable to verify the specific cause between HSW and CVD. Although many covariates were controlled and adjusted, there may still be potential and neglected confounders, like dietary patterns. Then, the HSW was only detected and calculated at a single point ^51^, which may trigger the misclassification. Nevertheless, this misclassification would lead to an underestimate but not an overestimation ^52^.

## 5. Conclusion

Higher HSW may be more likely to develop CVD. And in CVD patients, higher HSW indicated a poor prognosis. And the predictive value is much higher than RDW and MCHC. Individuals with females, aged <= 60 years, and DM have a higher risk of CVD when HSW increases. In conclusion, HSW is an innovative independent risk factor for CVD prevalence and long-term prognosis.

## Data Availability

All data produced in the present study are available upon reasonable request to the authors

## Abbreviations

ALB: albumin
ALT: alanine transaminase
BUN: blood urea nitrogen
Ca: total calcium
Cl: chloride
Fe: iron
GGT: gammaglutamyl transferase
GLB: and globulin
Glu: glucose
HCO3: bicarbonate
MCV: mean corpuscular volume
MPV: mean platelet volume
Na: sodium
RBC: red blood cells
TC: total cholesterol
TG: triglycerides
TP: total protein
UA: uric acid
BaP: basophils percentage
CIs: confidence intervals
CKD: chronic kidney disease
COPD: chronic obstructive pulmonary disease
CVD: Cardiovascular disease
DM: diabetes mellitus
eGFR: estimated glomerular filtration rate
Eo: eosinophils number count
EoP: eosinophils percentage
EV-SD: erythrocyte volume’s standard deviation
Hb: hemoglobin
Hem: hematocrit
HR: hazard ratios
HSW: Hemoglobin specific volume width
HSWQ: HSW’s quartiles
KM: Kaplan–Meier
MCH: mean cell hemoglobin
MCHC: mean corpuscular haemoglobin concentration
MCQ: medical conditions questionnaire
Mon: monocyte number
MonP: monocyte percentage
NHANES: national health and nutrition examination survey
OR: odds ratio
Plt: platelet count count
RCS: restricted cubic spline
RDW: red blood distribution width
RAR: red blood distribution width/serum albumin ratio
SegneP: segmented neutrophils percentage.

## Declarations

### Ethics approval and consent to participate

The NHANES protocol was approved by the Ethics Review Committee of the National Center for Health Statistics.

### Consent for publication

All patients/participants includec in this study provided their written informed consent.

### Availability of data and materials

NHANES data were derived from https://www.cdc.gov/nchs/nhanes/index.htm. And the mortality data were linked to https://www.cdc.gov/nchs/ndi/index.htm.

### Author’s contributions

LZ and JC wrote the original draft. LZ, JC, KYW, YL, and LMW performed the research. LZ, YZ, ZFF, and LLZ analyzed the data. XMZ, ZFF, LFH, and XMG designed the experiment and revised the manuscript.

## Acknowledgements

Thanks to Lifeng Han for proposing the idea of the article and Xiumei Gao for optimizing the framework of the article. Thanks to the NHANES.

