## Supplementary material for "Hemoglobin specific volume width might promote the prevalence and poor long-term prognosis of American cardiovascular disease adult patients: the NHANES 1999-2020": Table S1, Table S2, Table S3, Table S4, Table S5.

**Table S1.** The univariate and adjusted Logistic result

| **Variables** | **Univariate Logistic regression** | | **Adjusted Logistic regression** | |
| --- | --- | --- | --- | --- |
|  | **OR (95% CI)** | **P_value** | **OR (95%CI )** | **P_value** |
| HSW | 2.06(1.95,2.19) | <0.0001 | 1.46 | <0.0001 |
| HSWQ |  |  |  |  |
| Q1 | ref | ref | ref | ref |
| Q2 | 1.47(1.33,1.62) | <0.0001 | 1.07(0.95,1.20) | 0.25 |
| Q3 | 2.26(2.04,2.49) | <0.0001 | 1.19(1.05,1.35) | 0.01 |
| Q4 | 3.70(3.34,4.09) | <0.0001 | 1.54(1.36,1.73) | <0.0001 |
| RDW | 1.31(1.29,1.34) | <0.0001 | 1.14(1.10,1.17) | <0.0001 |
| MCHC | 0.16(0.11,0.23) | <0.0001 | 0.50(0.27,0.91) | 0.02 |
| Age | 1.08(1.07,1.08) | <0.0001 | 1.05(1.05,1.06) | <0.0001 |
| Sex |  |  |  |  |
| Female | ref | ref | ref | ref |
| Male | 1.27(1.18,1.36) | <0.0001 | 1.24(1.11,1.38) | <0.001 |
| CKD |  |  |  |  |
| no | ref | ref | ref | ref |
| yes | 5.00(4.67,5.35) | <0.0001 | 1.31(1.20,1.44) | <0.0001 |
| COPD |  |  |  |  |
| no | ref | ref | ref | ref |
| yes | 4.65(4.06,5.32) | <0.0001 | 2.26(1.92,2.65) | <0.0001 |
| Diabetes state |  |  |  |  |
| Health | ref | ref | ref | ref |
| preDM | 1.93(1.66,2.24) | <0.0001 | 1.05(0.89,1.24) | 0.56 |
| DM | 4.86(4.49,5.27) | <0.0001 | 1.58(1.40,1.78) | <0.0001 |
| Hypertension | |  |  |  |
| no | ref | ref | ref | ref |
| yes | 5.83(5.35,6.36) | <0.0001 | 1.95(1.75,2.18) | <0.0001 |
| Ethnicity |  |  |  |  |
| Mexican American | ref | ref | ref | ref |
| Non-Hispanic Black | 2.12(1.85,2.42) | <0.0001 | 1.30(1.09,1.56) | 0.004 |
| Non-Hispanic White | 2.34(2.07,2.65) | <0.0001 | 1.33(1.13,1.56) | <0.001 |
| Other Hispanic | 1.24(1.06,1.46) | 0.01 | 1.04(0.86,1.26) | 0.68 |
| Other Race | 1.76(1.42,2.17) | <0.0001 | 1.31(1.04,1.66) | 0.02 |
| WBC | 1.03(1.02,1.05) | <0.0001 | 1.01(0.96,1.06) | 0.73 |
| LymP | 0.96(0.95,0.96) | <0.0001 | 0.94(0.87,1.02) | 0.14 |
| MonP | 1.10(1.08,1.12) | <0.0001 | 0.96(0.88,1.05) | 0.39 |
| SegneP | 1.02(1.02,1.03) | <0.0001 | 0.94(0.87,1.02) | 0.17 |
| EoP | 1.07(1.06,1.08) | <0.0001 | 0.95(0.87,1.04) | 0.29 |
| BaP | 1.13(1.06,1.21) | <0.001 | 0.91(0.82,1.02) | 0.12 |
| Mon | 3.40(2.70,4.29) | <0.0001 | 1.07(0.58,1.97) | 0.84 |
| SeneP | 1.08(1.06,1.10) | <0.0001 | 1.03(0.96,1.10) | 0.44 |
| Eo | 2.46(2.11,2.86) | <0.0001 | 1.34(0.70,2.59) | 0.38 |
| RBC | 0.57(0.52,0.61) | <0.0001 | 0.89(0.32,2.45) | 0.82 |
| Hem | 0.96(0.95,0.97) | <0.0001 | 1.01(0.91,1.13) | 0.81 |
| MCV | 1.04(1.04,1.05) | <0.0001 | 1.02(0.97,1.07) | 0.52 |
| Plt | 1.00(0.99,1.00) | <0.0001 | 1.00(1.00,1.00) | <0.0001 |
| MPV | 1.07(1.02,1.12) | 0.01 | 0.99(0.93,1.05) | 0.76 |
| ALB | 0.33(0.30,0.36) | <0.0001 | 0.80(0.31,2.09) | 0.65 |
| ALT | 0.99(0.99,1.00) | <0.001 | 1.00(1.00,1.00) | 0.23 |
| BUN | 1.11(1.10,1.11) | <0.0001 | 1.01(1.00,1.01) | 0.16 |
| Ca | 0.89(0.80,0.99) | 0.03 | 1.11(0.98,1.26) | 0.10 |
| TC | 0.99(0.99,0.99) | <0.0001 | 0.99(0.99,0.99) | <0.0001 |
| HCO3 | 1.05(1.03,1.07) | <0.0001 | 0.97(0.95,0.99) | 0.01 |
| GGT | 1.00(1.00,1.00) | <0.0001 | 1.00(1.00,1.00) | 0.02 |
| Glu | 1.01(1.01,1.01) | <0.0001 | 1.00(1.00,1.00) | 0.53 |
| Fe | 0.99(0.99,1.00) | <0.0001 | 1.00(1.00,1.00) | 0.01 |
| TP | 0.63(0.58,0.68) | <0.0001 | 0.80(0.31,2.06) | 0.64 |
| TG | 1.00(1.00,1.00) | <0.0001 | 1.00(1.00,1.00) | <0.0001 |
| UA | 1.29(1.26,1.32) | <0.0001 | 1.07(1.03,1.11) | <0.001 |
| Na | 1.02(1.00,1.04) | 0.04 | 1.01(0.98,1.03) | 0.57 |
| Cl | 0.95(0.93,0.96) | <0.0001 | 0.99(0.97,1.01) | 0.24 |
| GLB | 1.26(1.17,1.35) | <0.0001 | 1.33(0.53,3.36) | 0.54 |

* The model was adjusted with age, sex, ethnicity, CKD, COPD, Diabetes, Hypertension, WBC, LymP, MonP, SegneP, EoP, BaP, Mon, SeneP, Eo, RBC, Hem, MCV, Plt, MPV, ALB, ALT, BUN, Ca, TC, HCO3, GGT, Glu, Fe, TP, TG, UA, Na, Cl, and GLB.

**Table S2.** The association of CVD in HSW with stratification of sex.

| **Sex** | **Variables** | **CVD** | | | | | | | |
| --- | --- | --- | --- | --- | --- | --- | --- | --- | --- |
|  |  | **OR (95% CI)^a^** | **P-value** | **OR (95% CI)^b^** | **P-value** | **OR (95% CI)^c^** | **P-value** | **OR (95% CI)^d^** | **P-value** |
| Male | HSW | 3.33(2.96,3.76) | <0.0001 | 1.71(1.52,1.92) | <0.0001 | 1.50(1.31,1.71) | <0.0001 | 1.37(1.18,1.60) | <0.0001 |
|  | Q1 | ref | ref | ref | ref | ref | ref | ref | ref |
|  | Q2 | 1.26(1.10,1.45) | 0.001 | 0.93(0.80,1.08) | 0.35 | 0.94(0.79,1.11) | 0.44 | 0.91(0.77,1.09) | 0.31 |
|  | Q3 | 2.26(1.97,2.59) | <0.0001 | 1.21(1.04,1.40) | 0.01 | 1.14(0.96,1.34) | 0.14 | 1.05(0.87,1.26) | 0.61 |
|  | Q4 | 4.41(3.82,5.08) | <0.0001 | 1.81(1.54,2.11) | <0.0001 | 1.56(1.30,1.86) | <0.0001 | 1.41(1.17,1.70) | <0.001 |
| Female | HSW | 1.75(1.63,1.88) | <0.0001 | 1.70(1.56,1.85) | <0.0001 | 1.47(1.34,1.62) | <0.0001 | 1.49(1.32, 1.68) | <0.0001 |
|  | Q1 | ref | ref | ref | ref | ref | ref | ref | ref |
|  | Q2 | 1.78(1.53,2.07) | <0.0001 | 1.35(1.15,1.58) | <0.001 | 1.28(1.06,1.54) | 0.01 | 1.28(1.07, 1.55) | 0.01 |
|  | Q3 | 2.30(1.99,2.67) | <0.0001 | 1.54(1.32,1.80) | <0.0001 | 1.42(1.20,1.68) | <0.0001 | 1.36(1.14, 1.62) | <0.001 |
|  | Q4 | 3.54(3.08,4.07) | <0.0001 | 2.29(1.98,2.64) | <0.0001 | 1.85(1.57,2.17) | <0.0001 | 1.67(1.40, 1.99) | <0.0001 |

a Model 1: unadjusted; b Model 2: adjusted with age; c Model 3: adjusted with age, ethnicity, CKD, COPD, Diabetes, and Hypertension; d Model 4: adjusted with age, ethnicity, CKD, COPD, Diabetes, Hypertension, WBC, LymP, MonP, SegneP, EoP, BaP, Mon, SeneP, Eo, RBC, Hem, MCV, Plt, MPV, ALB, ALT, BUN, Ca, TC, HCO3, GGT, Glu, Fe, TP, TG, UA, Na, Cl, and GLB.

**Table S3.** The association of CVD in HSW with stratification of age.

| **Age** | **Variables** | **CVD** | | | | | | | |
| --- | --- | --- | --- | --- | --- | --- | --- | --- | --- |
|  |  | **OR (95% CI)^a^** | **P-value** | **OR (95% CI)^b^** | **P-value** | **OR (95% CI)^c^** | **P-value** | **OR (95% CI)^d^** | **P-value** |
| >60 | HSW | 1.84(1.68,2.03) | <0.0001 | 1.87(1.69,2.06) | <0.0001 | 1.61(1.43,1.80) | <0.0001 | 1.35(1.18, 1.53) | <0.0001 |
|  | Q1 | ref | ref | ref | ref | ref | ref | ref | ref |
|  | Q2 | 1.08(0.94,1.24) | 0.29 | 1.08(0.94,1.25) | 0.27 | 1.06(0.91,1.24) | 0.46 | 1.00(0.85,1.17) | 0.99 |
|  | Q3 | 1.30(1.13,1.51) | <0.001 | 1.28(1.10,1.49) | 0.001 | 1.22(1.04,1.44) | 0.02 | 1.10(0.94,1.30) | 0.24 |
|  | Q4 | 2.01(1.77,2.28) | <0.0001 | 2.03(1.78,2.32) | <0.0001 | 1.75(1.51,2.02) | <0.0001 | 1.40(1.19,1.64) | <0.0001 |
| <=60 | HSW | 1.74(1.60,1.89) | <0.0001 | 1.81(1.65,1.98) | <0.0001 | 1.51(1.36,1.67) | <0.0001 | 1.57(1.35, 1.82) | <0.0001 |
|  | Q1 | ref | ref | ref | ref | ref | ref | ref | ref |
|  | Q2 | 1.34(1.12,1.61) | 0.002 | 1.33(1.11,1.60) | 0.002 | 1.23(1.02,1.50) | 0.03 | 1.12(0.92,1.36) | 0.26 |
|  | Q3 | 1.88(1.56,2.26) | <0.0001 | 1.89(1.58,2.28) | <0.0001 | 1.50(1.22,1.85) | <0.001 | 1.22(0.98,1.53) | 0.08 |
|  | Q4 | 2.73(2.32,3.21) | <0.0001 | 2.87(2.43,3.37) | <0.0001 | 1.90(1.59,2.27) | <0.0001 | 1.63(1.34,1.98) | <0.0001 |

a Model 1: unadjusted; b Model 2: adjusted with sex; c Model 3: adjusted with sex, ethnicity, CKD, COPD, Diabetes, and Hypertension; d Model 4: adjusted with sex, ethnicity, CKD, COPD, Diabetes, Hypertension, WBC, LymP, MonP, SegneP, EoP, BaP, Mon, SeneP, Eo, RBC, Hem, MCV, Plt, MPV, ALB, ALT, BUN, Ca, TC, HCO3, GGT, Glu, Fe, TP, TG, UA, Na, Cl, and GLB.

**Table S4.** The association of CVD in HSW with stratification of diabetes states.

| **Diabetes state** | **Variables** | **CVD** | | | | | | | |
| --- | --- | --- | --- | --- | --- | --- | --- | --- | --- |
|  |  | **OR (95% CI)^a^** | **P-value** | **OR (95% CI)^b^** | **P-value** | **OR (95% CI)^c^** | **P-value** | **OR (95% CI)^d^** | **P-value** |
| Health | HSW | 1.84(1.71,1.97) | <0.0001 | 1.55(1.41,1.70) | <0.0001 | 1.47(1.31,1.64) | <0.0001 | 1.45(1.27,1.65) | <0.0001 |
|  | Q1 | ref | ref | ref | ref | ref | ref | ref | ref |
|  | Q2 | 1.47(1.30,1.66) | <0.0001 | 1.10(0.97,1.24) | 0.13 | 1.11(0.97,1.26) | 0.12 | 1.12(0.98,1.28) | 0.09 |
|  | Q3 | 2.06(1.83,2.32) | <0.0001 | 1.23(1.09,1.39) | <0.001 | 1.20(1.05,1.38) | 0.01 | 1.18(1.01,1.37) | 0.03 |
|  | Q4 | 3.03(2.66,3.45) | <0.0001 | 1.68(1.47,1.92) | <0.0001 | 1.58(1.37,1.83) | <0.0001 | 1.49(1.27,1.76) | <0.0001 |
| preDM | HSW | 1.72(1.35,2.19) | <0.0001 | 1.52(1.17,1.99) | 0.002 | 1.41(1.05,1.90) | 0.02 | 1.29(0.94,1.77) | 0.11 |
|  | Q1 | ref | ref | ref | ref | ref | ref | ref | ref |
|  | Q2 | 0.96(0.69,1.34) | 0.83 | 0.72(0.51,1.02) | 0.06 | 0.75(0.52,1.07) | 0.11 | 0.76(0.52,1.11) | 0.15 |
|  | Q3 | 1.35(0.91,2.02) | 0.14 | 0.95(0.63,1.44) | 0.82 | 0.91(0.58,1.42) | 0.68 | 0.83(0.52,1.32) | 0.43 |
|  | Q4 | 2.24(1.56,3.22) | <0.0001 | 1.50(1.03,2.20) | 0.04 | 1.43(0.93,2.19) | 0.10 | 1.22(0.82,1.81) | 0.33 |
| DM | HSW | 1.92(1.70,2.16) | <0.0001 | 1.82(1.60,2.07) | <0.0001 | 1.68(1.44,1.96) | <0.0001 | 1.54(1.30,1.84) | <0.0001 |
|  | Q1 | ref | ref | ref | ref | ref | ref | ref | ref |
|  | Q2 | 1.33(1.06,1.65) | 0.01 | 1.24(0.98,1.56) | 0.08 | 1.17(0.91,1.50) | 0.23 | 1.12(0.86, 1.46) | 0.41 |
|  | Q3 | 1.96(1.63,2.37) | <0.0001 | 1.66(1.35,2.04) | <0.0001 | 1.58(1.27,1.97) | <0.0001 | 1.41(1.12, 1.78) | 0.004 |
|  | Q4 | 2.84(2.31,3.48) | <0.0001 | 2.35(1.88,2.92) | <0.0001 | 2.11(1.66,2.69) | <0.0001 | 1.77(1.36, 2.29) | <0.0001 |

a Model 1: unadjusted; b Model 2: adjusted with age and sex; c Model 3: adjusted with age and sex, ethnicity, CKD, COPD, and Hypertension; d Model 4: adjusted with age, sex, ethnicity, CKD, COPD, Hypertension, WBC, LymP, MonP, SegneP, EoP, BaP, Mon, SeneP, Eo, RBC, Hem, MCV, Plt, MPV, ALB, ALT, BUN, Ca, TC, HCO3, GGT, Glu, Fe, TP, TG, UA, Na, Cl, and GLB.

**Table S5.** The univariate and adjusted Cox result

|  | **Univariate Cox regression** | | **Adjusted Cox* regression** | |
| --- | --- | --- | --- | --- |
| **Variables** | **HR (95% CI)** | **P_value** | **HR (95%CI )** | **P_value** |
| HSW | 1.87(1.70,2.06) | <0.0001 | 1.70(1.48,1.95) | <0.0001 |
| HSWQ |  |  |  |  |
| Q1 | ref | ref | ref | ref |
| Q2 | 1.40(1.19,1.65) | <0.0001 | 1.18(0.99, 1.40) | 0.06 |
| Q3 | 1.75(1.46,2.08) | <0.0001 | 1.30(1.09, 1.55) | 0.004 |
| Q4 | 2.91(2.47,3.41) | <0.0001 | 1.88(1.53, 2.31) | <0.0001 |
| RDW | 1.23(1.19,1.28) | <0.0001 | 1.17(1.12,1.23) | <0.0001 |
| MCHC | 0.16(0.08,0.30) | <0.0001 | 0.19(0.09, 0.41) | <0.0001 |
| Age | 1.08(1.07,1.08) | <0.0001 | 1.06(1.05,1.07) | <0.0001 |
| Sex |  |  |  |  |
| Female | ref | ref | ref | ref |
| Male | 0.95(0.86,1.05) | 0.28 | 1.23(1.06,1.43) | 0.01 |
| CKD |  |  |  |  |
| no | ref | ref | ref | ref |
| yes | 3.16(2.83,3.53) | <0.0001 | 1.30(1.15,1.48) | <0.0001 |
| COPD |  |  |  |  |
| no | ref | ref | ref | ref |
| yes | 1.54(1.34,1.77) | <0.0001 | 1.40(1.21,1.63) | <0.0001 |
| Diabetes state |  |  |  |  |
| Health | ref | ref | ref | ref |
| preDM | 1.31(1.11,1.54) | 0.001 | 1.13(0.95,1.35) | 0.17 |
| DM | 1.65(1.45,1.87) | <0.0001 | 1.23(1.07,1.41) | 0.003 |
| Hypertension | |  |  |  |
| no | ref | ref | ref | ref |
| yes | 1.56(1.38,1.77) | <0.0001 | 1.00(0.88,1.13) | 0.95 |
| Ethnicity |  |  |  |  |
| Mexican American | ref | ref | ref | ref |
| Non-Hispanic Black | 1.33(1.07,1.65) | 0.01 | 0.94(0.74,1.20) | 0.64 |
| Non-Hispanic White | 1.66(1.35,2.05) | <0.0001 | 1.14(0.94,1.39) | 0.19 |
| Other Hispanic | 1.06(0.74,1.52) | 0.75 | 0.90(0.63,1.30) | 0.58 |
| Other Race | 1.07(0.70,1.65) | 0.74 | 0.82(0.55,1.23) | 0.34 |
| WBC | 1.03(1.02,1.04) | <0.0001 | 1.00(0.98,1.01) | 0.86 |
| LymP | 0.96(0.95,0.97) | <0.0001 | 0.99(0.95,1.03) | 0.68 |
| MonP | 1.05(1.03,1.07) | <0.0001 | 0.97(0.91,1.03) | 0.3 |
| SegneP | 1.03(1.02,1.04) | <0.0001 | 1.00(0.96,1.04) | 0.94 |
| EoP | 1.00(0.98,1.02) | 0.96 | 1.01(0.94,1.08) | 0.82 |
| BaP | 0.93(0.84,1.04) | 0.19 | 1.05(0.92,1.19) | 0.48 |
| Mon | 2.45(1.95,3.07) | <0.0001 | 1.81(1.09,3.01) | 0.02 |
| SeneP | 1.12(1.08,1.15) | <0.0001 | 1.06(0.99,1.14) | 0.12 |
| Eo | 1.14(0.94,1.39) | 0.18 | 0.78(0.42,1.46) | 0.44 |
| RBC | 0.49(0.44,0.55) | <0.0001 | 1.32(0.38,4.57) | 0.66 |
| Hem | 0.94(0.93,0.95) | <0.0001 | 0.95(0.83,1.09) | 0.47 |
| MCV | 1.04(1.03,1.05) | <0.0001 | 1.05(0.99,1.12) | 0.09 |
| Plt | 1.00(1.00,1.00) | <0.0001 | 1.00(1.00,1.00) | 0.01 |
| MPV | 1.00(0.95,1.05) | 0.88 | 0.94(0.89,0.99) | 0.01 |
| ALB | 0.40(0.34,0.47) | <0.0001 | 0.35(0.06,1.91) | 0.23 |
| ALT | 0.98(0.97,0.99) | 0.01 | 0.99(0.99,1.00) | 0.27 |
| BUN | 1.05(1.04,1.05) | <0.0001 | 1.01(1.01,1.02) | <0.001 |
| Ca | 0.81(0.69,0.96) | 0.01 | 0.91(0.77,1.07) | 0.23 |
| TC | 1.00(1.00,1.00) | 0.05 | 1.00(1.00,1.00) | 0.001 |
| HCO3 | 1.05(1.03,1.08) | <0.0001 | 1.00(0.97,1.03) | 0.83 |
| GGT | 1.00(1.00,1.00) | <0.001 | 1.00(1.00,1.00) | <0.0001 |
| Glu | 1.00(1.00,1.00) | <0.0001 | 1.00(1.00,1.00) | 0.29 |
| Fe | 0.99(0.99,1.00) | <0.0001 | 1.00(1.00,1.00) | 0.12 |
| TP | 0.90(0.80,1.01) | 0.08 | 1.87(0.35,9.94) | 0.46 |
| TG | 1.00(1.00,1.00) | 0.18 | 1.00(1.00,1.00) | 0.5 |
| UA | 1.15(1.12,1.19) | <0.0001 | 1.02(0.98,1.05) | 0.4 |
| Na | 0.98(0.96,1.00) | 0.08 | 1.01(0.98,1.04) | 0.55 |
| Cl | 0.94(0.93,0.95) | <0.0001 | 0.96(0.93,0.99) | 0.004 |
| GLB | 1.38(1.24,1.53) | <0.0001 | 0.67(0.13,3.49) | 0.63 |

* The model was adjusted with age, sex, ethnicity, CKD, COPD, Diabetes, Hypertension, WBC, LymP, MonP, SegneP, EoP, BaP, Mon, SeneP, Eo, RBC, Hem, MCV, Plt, MPV, ALB, ALT, BUN, Ca, TC, HCO3, GGT, Glu, Fe, TP, TG, UA, Na, Cl, and GLB.
